# Isolated Systolic and Diastolic Hypertension by the 2017 ACC/AHA Guidelines and Risk of Cardiovascular Disease: A Large Prospective Cohort Study

**DOI:** 10.1101/2020.04.19.20071530

**Authors:** Xian-Bo Wu, Fu-Rong Li

## Abstract

**IMPORTANCE:** The 2017 American College of Cardiology/American Heart Association (ACC/AHA) blood pressure (BP) guidelines lowered the hypertension threshold from a systolic blood pressure/diastolic blood pressure level of ≥140/90 mm Hg to ≥130/80 mm Hg. The cardiovascular impact of isolated systolic hypertension (ISH) and isolated diastolic hypertension (IDH) under the new definition remains unclear.

**OBJECTIVE:** To examine the associations of ISH and IDH defined by the 2017 ACC/AHA guidelines with risk of cardiovascular disease (CVD) among the UK population.

**DESIGN, SETTING, AND PARTICIPANTS:** We used data from the UK Biobank study, which is a prospective population-based cohort study. Participants were categorized into 5 groups: normal BP, normal high BP, ISH, IDH and systolic and diastolic hypertension (SDH).

**MAIN OUTCOMES AND MEASURES:** The primary endpoint for this study was the composite of nonfatal myocardial infarction (MI), nonfatal ischemic stroke (IS), nonfatal haemorrhagic stroke (HS) and CVD death. We also explored the results for the above mentioned CVD outcomes separately. Baseline BP measurements were obtained twice after the participant had been at rest for at least 5 minutes in a seated position.

**RESULTS:** We included 470,625 participants who were free of CVD at baseline and had available data on BP measures. During a median follow-up of 8.1 years, 13,157 CVD events were recorded, including 6,865 nonfatal MI, 3,415 nonfatal ISs, 1,118 nonfatal HSs, and 2,971 CVD deaths. According to the hypertension threshold of ≥130/80 mm Hg by the ACC/AHA guidelines, both ISH (HR 1.35, 95% CI 1.24-1.46) and IDH (HR 1.22, 95% CI 1.11-1.36) were significantly associated with a higher overall CVD risk as compared with normal BP. ISH was associated with most CVD risk, except for IS, while the excess CVD risk associated with IDH appeared to be driven mainly by MI. We found heterogeneity by sex and age regarding the effects of IDH on overall CVD risk, with significant associations in younger adults (age < 60 years) and women and null associations in men and older adults (age ≥60 years).

**CONCLUSIONS AND RELEVANCE:** Both ISH and IDH were associated with an increased risk of CVD among the UK population according to the ACC/AHA BP guidelines. Further research is needed to identify participants with IDH who have a particularly risk for developing CVD.

**Key points:** 

**Question:** What are the associations of isolated systolic hypertension (ISH) and isolated diastolic hypertension (IDH) by the 2017 ACC/AHA guidelines with CVD risk among the UK population?

**Findings:** In this prospective population-based cohort study of 470,625 UK participants, we found that compared with normotensive participants, those with ISH or IDH both had higher risk of overall CVD outcome. However, for participants with IDH, most CVD risk was driven by myocardial infarction and significant associations were observed in younger adults (age < 60 years) and women only.

**Meaning:** Our results indicate that by using the lower hypertension threshold by the 2017 ACC/AHA guidelines, both ISH and IDH were linked to a higher risk of CVD. Yet, results varied across age and sex for IDH, suggesting that more research is needed to identify participants with IDH who are at especially greater risk for developing CVD.

---

Hypertension can be diagnosed based on singular and combined elevation of systolic blood pressure (SBP) and diastolic blood pressure (DBP). People who fulfil only the SBP or only the DBP hypertension criteria are categorized as having isolated systolic hypertension (ISH) or isolated diastolic hypertension (IDH), respectively ^1^.

In 2017, the American College of Cardiology (ACC)/American Heart Association (AHA) lowered the hypertension criterion from an SBP/DBP level of ≥140/90 mm Hg to a threshold of ≥130/80 mm Hg ^2^. Since the release of the new guidelines, a number of studies have explored the change in prevalent hypertension in different populations ^3-6^ or cardiovascular disease (CVD) risk associated with the newly defined stage 1 hypertension (130-139/80-89 mm Hg) ^7 8^. To date, however, very few studies have focused on the impact of hypertension subtypes (e.g., ISH, IDH) defined by the ACC/AHA guidelines on CVD risk. A recent study of the US outpatient population reported that both systolic and diastolic hypertension independently predicted CVD events ^9^, regardless of BP cutoffs of 130/80 mmHg or 140/90 mmHg. However, this study did not explore the associations between hypertension subtypes and CVD risk. On the other hand, a study of multiple cohorts of US adults tried to clarify the associations between IDH and CVD risk by either hypertension definition ^10^, but the results for ISH were not available in that study.

Indeed, evidence regarding the associations of the redefined ISH and IDH by the 2017 ACC/AHA guidelines with CVD risk is limited, and the conclusions remained unclear. To fill the gap in the previous research, we used data from the UK Biobank to explore the associations of ISH and IDH with risk of CVD, using the BP threshold recommended by the 2017 ACC/AHA guidelines.

## Methods

### Study Population

Data for this study were derived from the UK Biobank study. The study design of the UK Biobank study has been previously reported in detail ^11, 12^. In brief, the UK Biobank study is a large prospective cohort study that recruited over 500,000 participants aged 40-69 years from 2006-2010 at baseline. It collected extensive phenotypic and genotypic data from 22 assessment centres across England, Wales, and Scotland, covering a variety of different socioeconomic and ethnic settings. In the present study, we excluded 1,243 individuals with missing values for SBP or DBP, 29,329 individuals with prevalent CVD (angina, myocardial infarction (MI) and stroke) at baseline and 1,240 individuals who were lost to follow-up. The final sample for analysis included 470,625 participants.

The UK Biobank has ethical approval from the Northwest Multi-Center Research Ethics Committee, and written informed consent was obtained from all participants.

### Assessment of Blood Pressure

Baseline BP measurements were obtained twice after the participant had been at rest for at least 5 minutes in a seated position using a digital BP monitor (Omron HEM-7015IT; OMRON Healthcare Europe B.V., Hoofddorp, Netherlands) with a suitably sized cuff. Mean SBP and DBP values were calculated from 2 automated (*n*=439,829) or 2 manual (*n*=30,796) BP measurements by trained nurses.

### Assessment of Outcomes

The primary endpoint for this study was the composite of nonfatal MI, nonfatal ischaemic stroke (IS), nonfatal haemorrhagic stroke (HS) and CVD death. We also explored the results for the above CVD outcomes separately. Information on CVD events and the timing of events was identified by linking to the Scottish Morbidity Records for participants from Scotland and health episode statistics for participants from England and Wales. The date and cause of death were identified by linking to the death registries of the National Health Service (NHS) Information Centre for participants from England and Wales and the NHS Central Register Scotland for participants from Scotland ^12^. At the time of analysis, hospital admission data were available up to March 29, 2017, and mortality data were available up to February 12, 2018 for England and Wales and August 12, 2016 for Scotland; therefore, these dates were used as the end of follow-up. Person-time was calculated from the date of baseline assessment to the date of diagnosis of the event, death, or the end of follow-up, whichever occurred first.

The International Classification of Diseases, tenth revision (ICD-10) codes were included in death records, while ICD-10 and International Classification of Diseases, ninth revision (ICD-9) codes were used in medical records. MI was defined as ICD-9 codes 410-414 and ICD-10 codes I20-I25. IS was defined as ICD-9 codes 433-434 and ICD-10 code I63; and HS was defined as ICD-9 codes 430-432 and ICD-10 codes I60-I62 ^13^. CVD death was defined as ICD-10 codes I00-I99.

### Assessment of Covariates

We used a baseline touch screen questionnaire to collect information on the following potential confounders: sociodemographic information (age, sex, ethnicity); area-based social deprivation (Townsend score, a composite measure of socioeconomic deprivation and household income); lifestyle (smoking, drinking, dietary habits and physical activity); and self-reported medical conditions (medications to treat high cholesterol and hypertension; history of diabetes and long-standing illness). A non-stretchable tape was used to measure height, and the Tanita BC-418 MA body analyser was used to measure weight ^14^. Body mass index (BMI) was calculated as weight in kilograms divided by height in metres squared (kg/m^2^). Circulating lipid profiles and serum cystatin C were also used for analysis. Long-standing illness was measured using the following question: “Do you have any long-standing illness, disability or infirmity?”. The ion-selective electrode method was used to measure sodium levels in stored urine samples. Dietary habits were assessed through a food frequency questionnaire. The healthy diet score was calculated using the medians of several dietary factors as follows ^13^: red meat intake less than three times each week; vegetable intake at least four tablespoons each day; fruit intake of at least three pieces each day; fish intake of at least four times each week; cereal intake of at least five bowls each week; and urinary sodium concentration (measured in stored urine samples using ion-selective electrode method ^15^) less than 68.3 mmol/L. Each favourable diet factor was assigned a score of 1 point, resulting in a total diet score ranging from 0 to 6. The estimated glomerular filtration rate (eGFR) was calculated by the Chronic Kidney Disease Epidemiology Collaboration (CKD–EPI) equation using serum cystatin C equations as previously reported ^16^. Physical activity was assessed by the International Physical Activity Questionnaire short form. We used the summed metabolic equivalents of energy (METs) per week for all activity in the analysis.

### Statistical Analysis

Continuous variables are presented as the means ± standard deviations (SDs), while categorical variables are presented as frequencies (%). Participants were stratified into 5 BP categories: 1) normal (SBP <120 mm Hg and DBP <80 mm Hg), 2) normal high (SBP 120 to 129 mm Hg and DBP <80 mm Hg), 3) ISH (SBP ≥130 mm Hg and DBP <80 mm Hg), 4) IDH (SBP <130 mm Hg and DBP ≥80 mm Hg), and 5) a combination of ISH and IDH (SDH) (SBP ≥130 mm Hg and DBP ≥80 mm Hg). Participants with normal BP were used as the reference. For supplementary analysis, we combined the normal BP group and normal high group, resulting in a new normal BP category as the reference (<130/80 mm Hg). To facilitate comparisons between guidelines, we also divided the sample into 5 categories by using the hypertension threshold (≥140/90 mm Hg) recommended by the National Institute of Clinical Excellence (NICE) guidelines in the UK.

Multivariable Cox regression models were used to calculate the hazard ratios (HRs) and 95% confidence intervals (CIs) for the associations between hypertension subtypes and CVD risk. Responses of “not known” or “prefer not to answer” to the covariates were considered missing values. Missing values accounted for <6% of all the covariates. Participants with missing values for any of the covariates were assigned to a separate “unknown” category. Two models were used to account for potential confounders. Model 1 included age (continuous) and sex. Model 2 further included ethnicity (white, mixed, Asian or Asian British, black or black British, Chinese, or other ethnic group), Townsend scores (in quintiles), BMI (in quintiles), smoking (never, previous, current), alcohol consumption (daily or almost daily, three or four times a week, once or twice a week, one to three times a month, special occasions only, never), healthy diet score, hypertensive medication use, cholesterol-lowering medication use, history of diabetes, eGFR and long-standing illness.

For the subgroup analysis, we examined the associations between BP categories and the primary outcome stratified by age (<60 years or ≥60 years) and sex. Several sensitivity analyses were performed to test the robustness of the primary result: 1) additional adjustment for physical activity; 2) additional adjustment for lipid profile, including serum concentrations of low-density lipoprotein cholesterol, high-density lipoprotein cholesterol and triglyceride; 3) combination of the aforementioned sensitivity analyses; 4) exclusion of those who had CVD events in the first two years of follow-up; 5) exclusion of those with missing variables; 6) exclusion of those taking antihypertensive medications; and 7) additional adjustment for baseline SBP or DBP for IDH or ISH, respectively.

Analyses were conducted using Stata version 14.0 (College Station, Texas). A *P*-value <0.05 was considered indicative of statistical significance.

## Results

### Baseline Characteristics

This study consisted of 470,625 participants (mean age 57 years), of whom 208,231 (44.3%) were men. The percentages of participants with ISH, IDH, and SDH were 15.9%, 7.5%, and 51.8% under the ACC/AHA guidelines, and 25.9%, 3.2%, and 21.3% under the NICE guidelines (Fig. 1).

**Figure 1.**
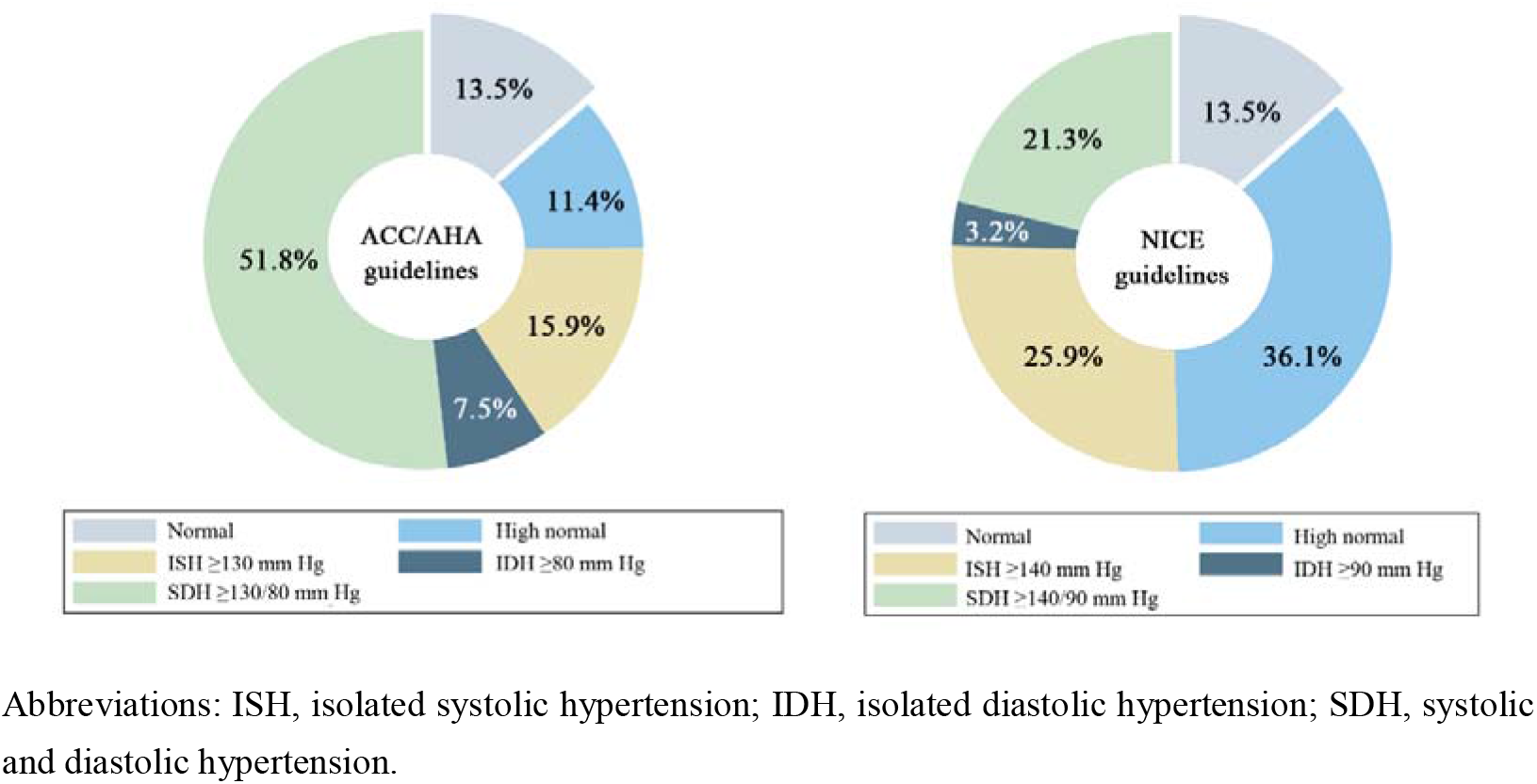
Participants meeting the definitions for hypertension subtypes by different guidelines.

Baseline characteristics of the study participants by normal BP status and hypertension subtypes are presented in Table 1. The mean age of the study participants with ISH was 59.3 years, while those with IDH had a mean age of 52.5 years. As compared to participants with normal BP, those with all other hypertension subtypes had a higher mean SBP, DBP and BMI. Moreover, hypertensive participants were more likely to have lower Townsend scores (except for IDH), take hypertensive and cholesterol-lowering medications, and have a history of diabetes or long-term illness, but they were less likely to be current smokers and have a lower eGFR level. With the exception of those with ISH, hypertensive participants also tended to have lower healthy diet scores.

**Table 1.** Baseline characteristics by normal BP status and hypertension subtype according to the ACC/AHA guidelines.

| Characteristics | Normal | High normal | ISH | IDH | SDH |
| --- | --- | --- | --- | --- | --- |
| No. of participants | 63,395 | 53,611 | 74,634 | 35,332 | 243,653 |
| Age, y | 52.1±7.8 | 54.6±8.1 | 59.3±7.5 | 52.5±7.6 | 57.3±7.7 |
| Men | 24.7 | 39.6 | 41.9 | 38.4 | 51.9 |
| BMI, kg/m<sup>2</sup> | 25.0±4.0 | 25.9±4.1 | 26.6±4.3 | 28.0±5.1 | 28.3±4.8 |
| SBP, mm Hg | 111.5±6.2 | 124.4±2.8 | 142.4±11.5 | 123.7±4.7 | 151.9±15.7 |
| DBP, mm Hg | 69.4±5.8 | 73.1±4.7 | 74.3±4.4 | 83.9±3.8 | 90.1±7.5 |
| Ethnicity |  |  |  |  |  |
| White | 92.7% | 94.3% | 95.9% | 92.1% | 94.3% |
| Mixed | 0.9% | 0.7% | 0.4% | 0.8% | 0.5% |
| Asian or Asian British | 2.4% | 1.8% | 1.4% | 2.7% | 1.8% |
| Black or black British | 1.6% | 1.4% | 1.0% | 2.3% | 1.8% |
| Chinese | 0.6% | 0.3% | 0.2% | 0.5% | 0.3% |
| Other ethnic group | 1.3% | 1.0% | 0.6% | 1.3% | 0.8% |
| Townsend score* | -1.1±3.2 | -1.3±3.1 | -1.5±3.0 | -1.1±3.2 | -1.4±3.0 |
| Alcohol drinking |  |  |  |  |  |
| Never | 9.0% | 8.2% | 8.5% | 8.2% | 7.1% |
| Special occasions only | 12.6% | 11.7% | 12.2% | 12.5% | 10.6% |
| One to three times a month | 13.4% | 11.9% | 11.2% | 12.1% | 10.3% |
| One or two times a week | 27.3% | 26.7% | 25.5% | 27.2% | 25.3% |
| Three or four times a week | 22.1% | 23.1% | 22.5% | 22.2% | 24.0% |
| Daily or almost daily | 15.4% | 18.3% | 19.9% | 17.7% | 22.6% |
| Smoking |  |  |  |  |  |
| Never | 57.2% | 57.0% | 54.5% | 56.6% | 54.9% |
| Former | 29.6% | 31.3% | 35.4% | 30.5% | 35.1% |
| Current | 12.8% | 11.3% | 9.6% | 12.4% | 9.5% |
| Healthy diet score | 3.0±1.4 | 3.0±1.4 | 3.1±1.4 | 2.8±1.4 | 2.8±1.4 |
| Hypertensive medication use | 2.7% | 4.3% | 9.7% | 7.3% | 14.1% |
| Cholesterol-lowering medication use | 6.1% | 9.8% | 18.1% | 9.5% | 15.5% |
| Diabetes | 3.3% | 4.8% | 8.9% | 4.8% | 6.6% |
| Long-term illness | 25.2% | 26.9% | 31.7% | 29.6% | 30.2% |
| eGFR, ml min<sup>-1</sup> per 1.73 m<sup>2</sup> | 94.2±15.2 | 91.5±15.6 | 86.4±16.2 | 92.0±15.8 | 87.3±15.6 |

Abbreviations: ISH, isolated systolic hypertension; IDH, isolated diastolic hypertension; SDH, systolic and diastolic hypertension. \*Positive values indicate areas with high material deprivation, whereas those with negative values indicate relative affluence.

### Associations between Hypertension Subtypes and Risk of CVD

During a median follow-up of 8.1 years, a total of 13,157 CVD events were recorded, including 6,865 nonfatal MI, 3,415 nonfatal IS, 1,118 nonfatal HS, and 2,971 CVD deaths. Using the hypertension cutoff of ≥130/80 mm Hg according to the ACC/AHA guideline, the multivariable-adjusted HRs (95% CI) of overall CVD outcome were 1.08 (0.98-1.18) for high normal BP, 1.35 (1.24-1.46) for ISH, 1.22 (1.11-1.36) for IDH, and 1.51 (1.40-1.62) for SDH, respectively, compared with those with normal BP (Figure 2). Regarding the associations between ISH and individual CVD outcomes (Figure 3), a null association was found for nonfatal IS (HR 1.15, 95% CI 0.98-1.34), whereas significant results were observed for nonfatal MI (HR 1.45, 95% CI 1.29-1.63), nonfatal HS (HR 1.53, 95% CI 1.18-2.00), and CVD mortality (HR 1.23, 95% CI 1.04-1.46). Moreover, for IDH, only the result for nonfatal MI was evident (HR 1.27, 95% CI 1.10-1.47). The results for SDH were consistently significant for all CVD events. For the NICE guideline (cutoff of ≥140/90 mm Hg) (Figure 2 and Supplemental Figure 1), compared with those with normal BP, all hypertension phenotypes were associated with higher risks of all CVDs, with the exception of CVD mortality, indicating a null association for IDH (HR 1.15, 95% CI 0.88-1.49). We analysed the association of BP as a continuous variable with the risk of CVD by different baseline BP cutoffs (Supplemental Table 1). SBP was consistently associated with higher risks of CVD regardless of DBP cutoffs (DBP <80 mm Hg or DBP <90 mm Hg). We observed significant results for overall CVD and MI associated with DBP as a continuous variable, but not for IS, HS or CVD death with the SBP cutoff of < 130 mm Hg; however, the risks for IS and HS became evident when using the SBP cutoff of < 140 mm Hg, even though the non-significant result for CVD death remained unchanged.

**Figure 2.**
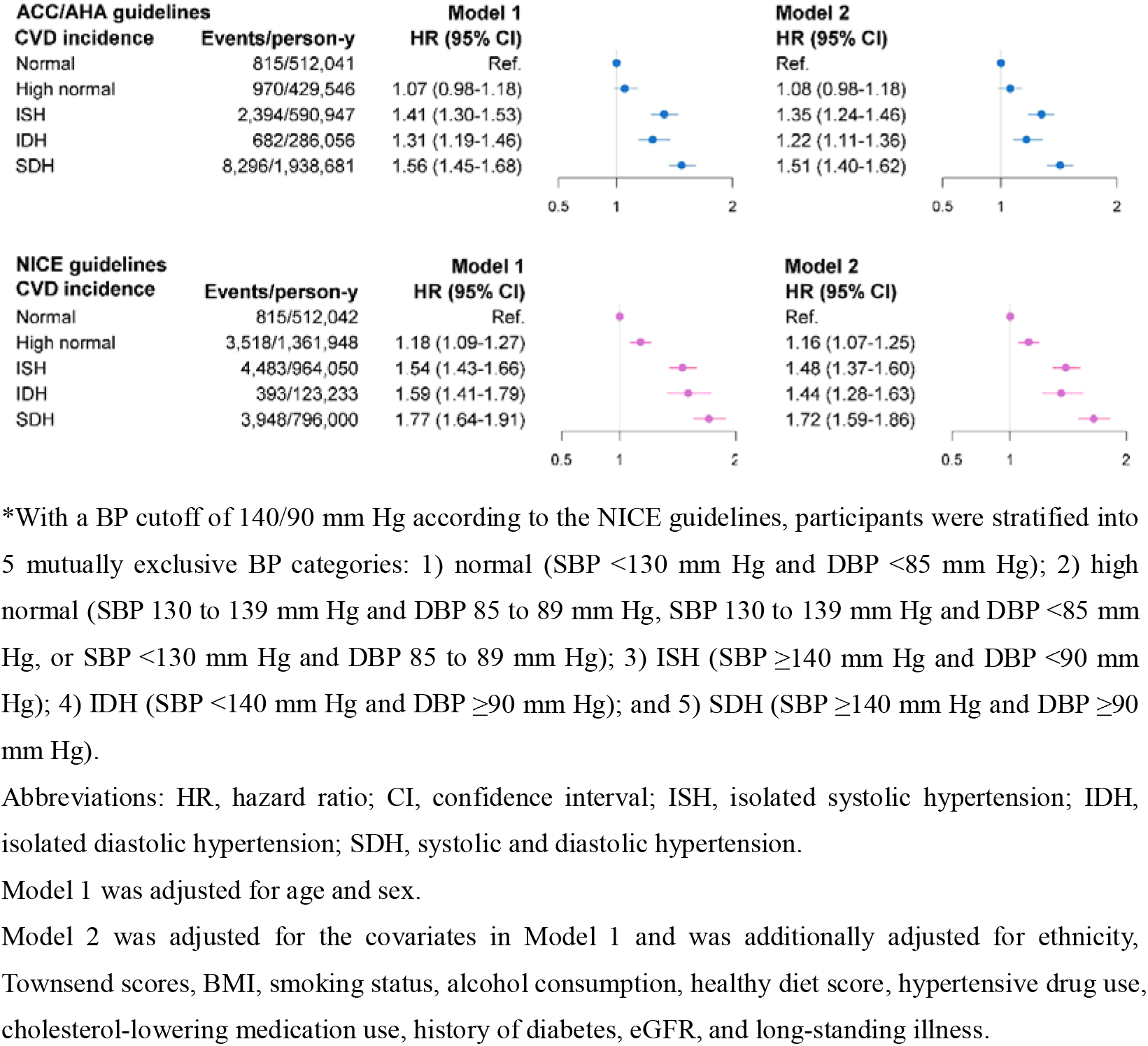
Associations of hypertension subtype according to the ACC/AHA guidelines and NICE guidelines with the risk of CVD.

**Figure 3.**
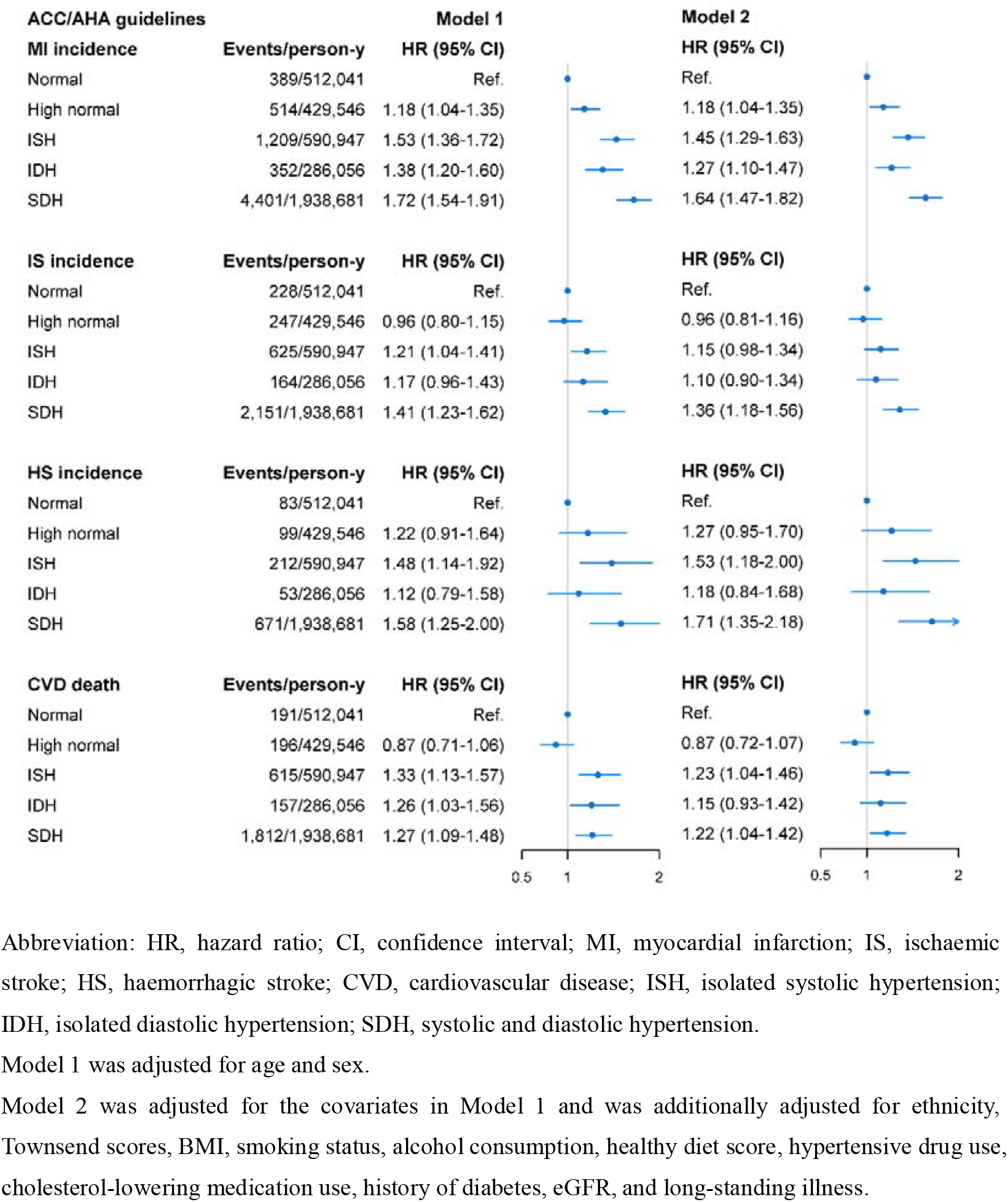
Associations of hypertension subtype according to the ACC/AHA guidelines with the risk of MI, IS, HS, and CVD death.

Subgroup analyses by age showed that the primary results were consistent for the young adults (aged<60 years), while for older adults (aged ≥60 years), the primary results remained similar for ISH and SDH but lost statistical significance for IDH (HR 1.08, 95% CI 0.94-1.28). Notably, we also found that men with IDH did not have a higher risk of CVD (HR 1.07, 95% CI 0.94-1.23), whereas the results were evident for all hypertension subtypes among women (Supplemental Figure 2).

### Exploratory Analysis and Sensitivity Analysis

Redefining the normal BP group by combining the original normal group and high normal group did not substantially change the results, except for the link between IDH and CVD death, which turned out to be statistically significant (HR 1.23, 95% CI 1.02-1.48) (Supplemental Figure 3). Sensitivity analysis yielded similar results after additional adjustment for physical activity, serum lipids, or a combination of the aforementioned analyses. Exclusion of participants with any missing covariates or those taking antihypertensive medications also yielded similar results (Supplemental Table 2). Of note, the significant results remained unchanged after additional adjustment for baseline SBP for IDH or baseline DBP for ISH, regardless of hypertension definition (Supplemental Table 3).

## Discussion

The present study evaluated the associations of different subtypes of hypertension, defined by the 2017 ACC/AHA guideline, with the risk of CVD outcomes in a large prospective cohort study of the UK population. We found that the excess overall CVD risk was evident among participants with ISH or IDH compared to their normotensive counterparts. The findings regarding ISH and IDH persisted across a series of sensitivity analyses, even after taking the baseline DBP and SBP into account. We found that the effects of IDH on CVD were stronger in women and younger adults (age <60 years) and null in men and older adults (age ≥60 years).

Few prospective studies have assessed the associations between ISH and IDH and CVD risk, especially in the context of the new definition according to the ACC/AHA guidelines. Flint *et al*. examined the impact of outpatient BP on CVD risk among 1.3 million US adults (median age 53 years) and found that both SBP and DBP independently predicted adverse CVD events, regardless of the definition of hypertension (≥140/90 mm Hg or ≥130/80 mm Hg) ^9^. Unfortunately, this study did not report the results for isolated hypertension as simple binary variables. Yano *et al*. reported that among 27,081 younger and middle-aged US adults (mean age, 34 years), all hypertension subtypes, defined by the threshold of ≥140/90 mm Hg, were associated with an increased risk of CVD ^17^; similar findings were also reported from a cohort study of 169,871 Chinese men and women (aged ≥40 years) ^18^. As most of the hypertension phenotypes were defined by the BP cutoff of ≥140/90 mm Hg in the above studies, their results may not be applicable to the current ACC/AHA guidelines.

Although a few studies have found positive results for IDH under the BP threshold of ≥140/90 mm Hg ^17, 18^, this was not the case in many previous studies. In fact, while the detrimental effects of ISH and SDH on CVD are without much controversy, the situation is less clear for IDH — the existing evidence regarding this issue is somewhat mixed. For instance, a recent study by McEvoy *et al*. failed to find significant results by either hypertension definition for IDH. In that study, a comprehensive analysis was performed to explore the associations of IDH with the risk of CVD among US adults from several cohort studies (ARIC, NHANES and CLUE II) ^10^. The uncertainty in the CVD risk related to IDH was also reflected by the null associations in other studies ^19-22^. The current study showed that by using the threshold of 130/80 mm Hg, ISH was a significant predictor of most CVD events, yet the excess CVD risk associated with IDH appeared to be driven mainly by MI; the associations between IDH and other CVD outcomes (IS, HS, CVD death) were not statistically significant. The lack of statistically significant associations of IDH with some of the CVD outcomes calls into question the pathogenicity of IDH.

Differences in biological processes may be responsible for the stronger effects of ISH than IDH on CVD risk ^23, 24^. ISH is characterized by higher stroke volume and/or aortic stiffness, and the occurrence of this phenotype is believed to reflect the high BP for multiple organs (e.g., brain, heart, and kidneys) ^25, 26^, which may result in poorer outcomes. Additionally, ISH is more likely to be the end point of SBP and DBP, both of which would be risk factors for CVD ^27^. IDH is a consequence of an increase in arteriolar resistance only ^21, 23^; as a result, participants with IDH may bear less vascular burden than those with ISH. In fact, a clear understanding of the pathophysiologic mechanisms underlying different hypertension phenotypes may help establish appropriate treatment recommendations in clinical practice.

An important finding of this study is that the increased risks of CVD associated with IDH were mainly concentrated in women or those aged <60 years. In general, women differ significantly from men in central arterial pressure and have higher vascular loading conditions ^28-30^; in addition, the association between arterial stiffness and left ventricular diastolic function appeared to be stronger in women than in men ^31^, which may collectively explain the sex-specific associations. A study of the UK Biobank also confirmed that BP was more strongly associated with MI in women than in men ^32^. On the other hand, previous studies have found that ISH is most common among elderly individuals, while IDH is more frequent in young and middle-aged persons ^1^. As the CVD risk increases by age, the age-specific prevalence of ISH and IDH may result in the differences in the CVD risks associated with these two phenotypes. In summary, these findings may indicate that tailored strategies for hypertension management would be needed based on age and sex.

### Strengths and Limitations

To the best of our knowledge, this is the first study to verify the validation of ISH and IDH according to the ACC/AHA guideline in predicting CVD risk. The strength of the study is that the cohort is large and has a median follow-up period of 8.1 years and 13,157 CVD events, which allow us to perform analyses by subtypes of hypertension and CVD events. A few limitations should also be acknowledged. First, BP measurements were collected only at a single visit, and changes over time would attenuate findings towards the null hypothesis, causing underestimation of the true associations. Second, the potential risk of bias due to “white-coat” effects should also be taken into account, as a patient’s BP measured in a clinic or office may be higher than their ambulatory pressure. Third, the study was based on a sample of the UK population, which limits the generalizability of the findings to other populations.

## Conclusions

Both ISH and IDH were associated with an increased risk of CVD among the UK population according to the ACC/AHA BP guidelines. Further research is needed to identify participants with IDH who have a particularly risk for developing CVD.

## Data Availability

Data were available in UK Biobank

https://www.ukbiobank.ac.uk/

---

**Supplemental Table 1.** Multivariate-adjusted HR (95% CIs) for BP as a continuous variable for the risk of CVD by different baseline BP cutoffs.

| Model | SBP, each 5 mm Hg increase |  | DBP, each 5 mm Hg increase |  |
| --- | --- | --- | --- | --- |
|  | DBP <80 mm Hg | DBP <90 mm Hg | SBP <130 mm Hg | SBP <140 mm Hg |
| CVD incidence |  |  |  |  |
| Model 1 | 1.07 (1.05-1.09) | 1.05 (1.04-1.05) | 1.06 (1.03-1.09) | 1.06 (1.04-1.08) |
| Model 2 | 1.05 (1.03-1.07) | 1.04 (1.04-1.05) | 1.03 (1.00-1.06) | 1.04 (1.02-1.05) |
| MI incidence |  |  |  |  |
| Model 1 | 1.06 (1.04-1.07) | 1.05 (1.04-1.06) | 1.08 (1.04-1.12) | 1.07 (1.04-1.09) |
| Model 2 | 1.05 (1.03-1.06) | 1.05 (1.04-1.06) | 1.04 (1.00-1.08) | 1.04 (1.01-1.06) |
| IS incidence |  |  |  |  |
| Model 1 | 1.04 (1.02-1.06) | 1.04 (1.03-1.05) | 1.02 (0.97-1.08) | 1.06 (1.02-1.09) |
| Model 2 | 1.03 (1.01-1.05) | 1.04 (1.02-1.05) | 1.00 (0.95-1.06) | 1.04 (1.00-1.07) |
| HS incidence |  |  |  |  |
| Model 1 | 1.05 (1.02-1.08) | 1.04 (1.02-1.06) | 1.03 (0.95-1.13) | 1.07 (1.01-1.13) |
| Model 2 | 1.05 (1.01-1.08) | 1.04 (1.02-1.07) | 1.05 (0.96-1.15) | 1.08 (1.02-1.15) |
| CVD death |  |  |  |  |
| Model 1 | 1.07 (1.05-1.09) | 1.04 (1.03-1.06) | 1.01 (0.95-1.07) | 1.02 (0.99-1.06) |
| Model 2 | 1.05 (1.03-1.07) | 1.04 (1.02-1.05) | 0.98 (0.93-1.04) | 1.00 (0.96-1.04) |

Abbreviation: HR, hazard ratio; CI, confidence interval; SBP, systolic blood pressure; DBP, diastolic blood pressure; MI, myocardial infarction; IS, ischaemic stroke; HS, haemorrhagic stroke; CVD, cardiovascular disease;
Model 1 was adjusted for age and sex.
Model 2 was adjusted for the covariates in Model 1 and was additionally adjusted for ethnicity, Townsend scores, BMI, smoking status, alcohol consumption, healthy diet score, hypertensive drug use, cholesterol-lowering medication use, history of diabetes, eGFR, and long-standing illness.

**Supplemental Table 2.** Sensitivity analyses of hypertension subtype by ACC/AHA definition and risk of CVD.

|  | Normal | High normal | ISH | IDH | SDH |
| --- | --- | --- | --- | --- | --- |
| Model 2 + physical activity |  |  |  |  |  |
| Multivariable adjusted (HR [95% CI]) | Ref. | 1.08 (0.98-1.18) | 1.34 (1.24-1.46) | 1.22 (1.11-1.36) | 1.50 (1.40-1.62) |
| Model 2 + lipid profile* |  |  |  |  |  |
| Multivariable adjusted (HR [95% CI]) | Ref. | 1.07 (0.98-1.18) | 1.34 (1.24-1.46) | 1.19 (1.07-1.32) | 1.48 (1.37-1.60) |
| Model 2 + physical activity and lipid profile |  |  |  |  |  |
| Multivariable adjusted (HR [95% CI]) | Ref. | 1.07 (0.98-1.18) | 1.34 (1.24-1.46) | 1.19 (1.07-1.32) | 1.48 (1.37-1.59) |
| Exclusion of missing variables |  |  |  |  |  |
| Multivariable adjusted (HR [95% CI]) | Ref. | 1.09 (0.99-1.20) | 1.35 (1.24-1.47) | 1.21 (1.09-1.35) | 1.51 (1.40-1.63) |
| Exclusion of those with incident CVD in the first 2 years of follow-up |  |  |  |  |  |
| Multivariable adjusted (HR [95% CI]) | Ref. | 1.13 (1.02-1.25) | 1.39 (1.27-1.52) | 1.23 (1.10-1.38) | 1.55 (1.43-1.69) |
| Exclusion of those taking antihypertensive medications |  |  |  |  |  |
| Multivariable adjusted (HR [95% CI]) | Ref. | 1.10 (1.00-1.21) | 1.39 (1.27-1.52) | 1.28 (1.15-1.43) | 1.62 (1.50-1.76) |

Abbreviations: HR, hazard ratio; CI, confidence interval; PA, physical activity; CVD, cardiovascular disease; ISH, isolated systolic hypertension; IDH, isolated diastolic hypertension; SDH, systolic and diastolic hypertension.
Model 2 was adjusted for age, sex, ethnicity, Townsend scores, BMI, smoking status, alcohol consumption, healthy diet score, hypertensive drug use, cholesterol-lowering medication use, history of diabetes, eGFR, and long-standing illness.
\*Lipid profile refers to the serum concentrations of low-density lipoprotein cholesterol, high-density lipoprotein cholesterol and triglyceride.

**Supplemental Table 3.** Associations of ISH and IDH with CVD risk by ACC/AHA definition, taking baseline BP into account.

|  | Normal | ISH | IDH |
| --- | --- | --- | --- |
| ACC/AHA guidelines<br>(HR [95% CI]) | Ref. | 1.28 (1.17-1.40) | 1.34 (1.16-1.55) |
| NICE guidelines<br>(HR [95% CI]) | Ref. | 1.41 (1.29-1.54) | 1.38 (1.17-1.62) |

Abbreviations: HR, hazard ratio; CI, confidence interval; ISH, isolated systolic hypertension; IDH, isolated diastolic hypertension; SDH, systolic and diastolic hypertension; BP, blood pressure.
Multifactorial adjustments were made for age, sex, ethnicity, Townsend scores, BMI, smoking status, alcohol consumption, healthy diet score, hypertensive drug use, cholesterol-lowering medication use, history of diabetes, eGFR, and long-standing illness.
For ISH, the multivariable-adjusted model was further adjusted for baseline DBP; whereas for IDH, the multivariable adjusted model was further adjusted for baseline SBP.

**Supplemental Figure 1.**
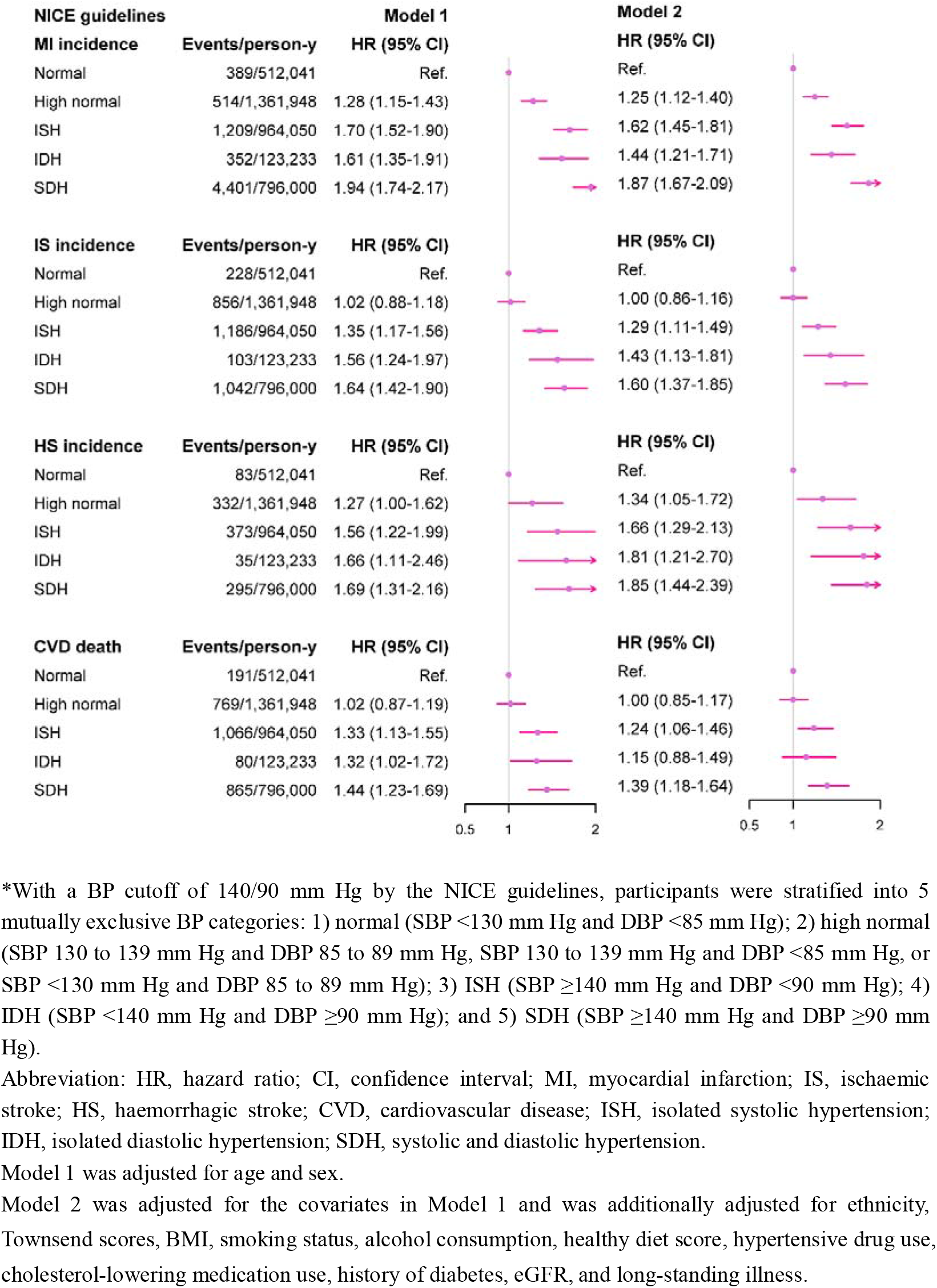
Multivariate-adjusted HRs (95% CIs) for the risk of CVD by hypertension subtype according to the NICE definition*.

**Supplemental Figure 2.**
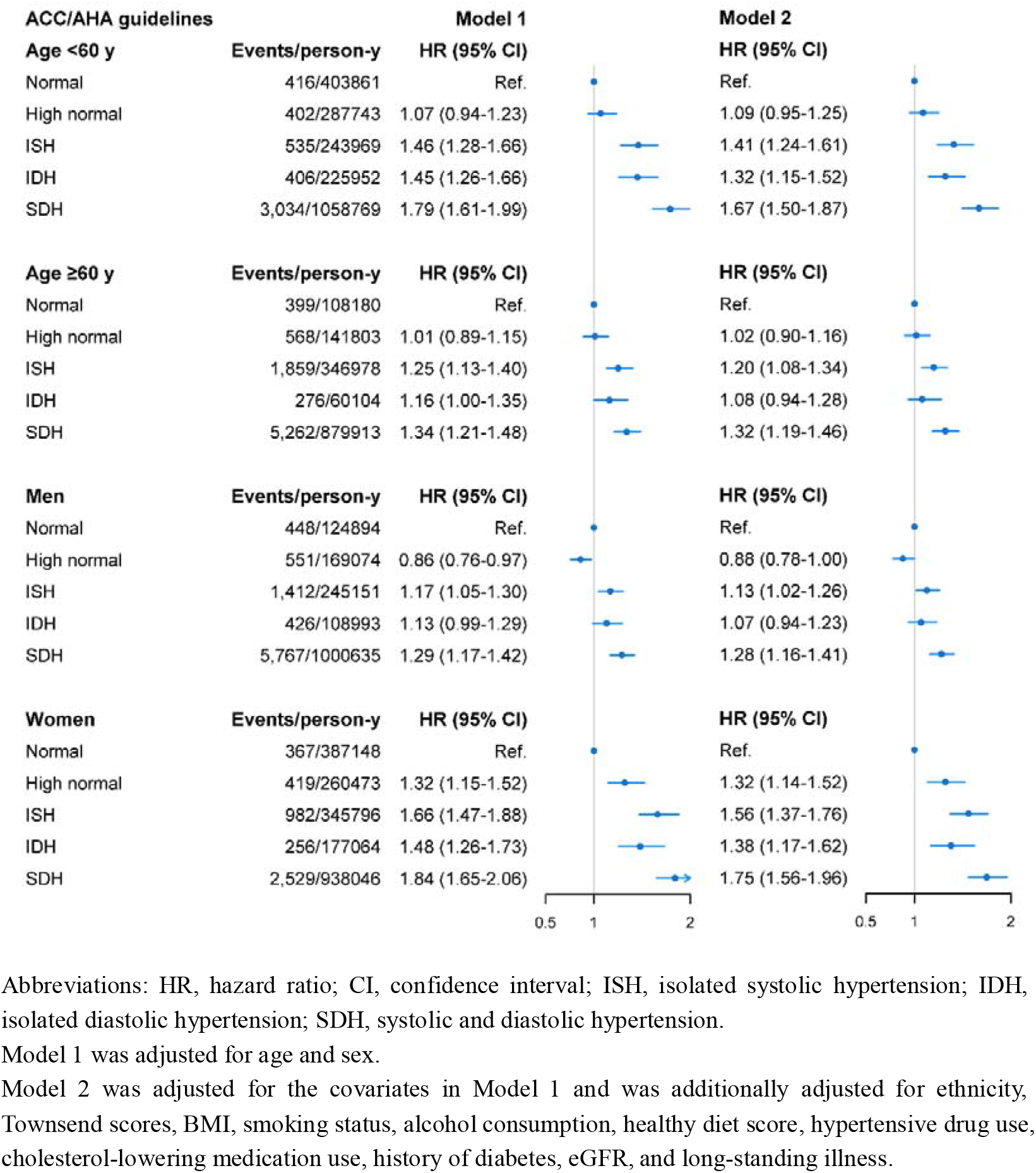
Age and sex-specific multivariate-adjusted HRs (95% CIs) for composite CVD outcome by hypertension subtype according to the ACC/AHA definition.

**Supplemental Figure 3.**
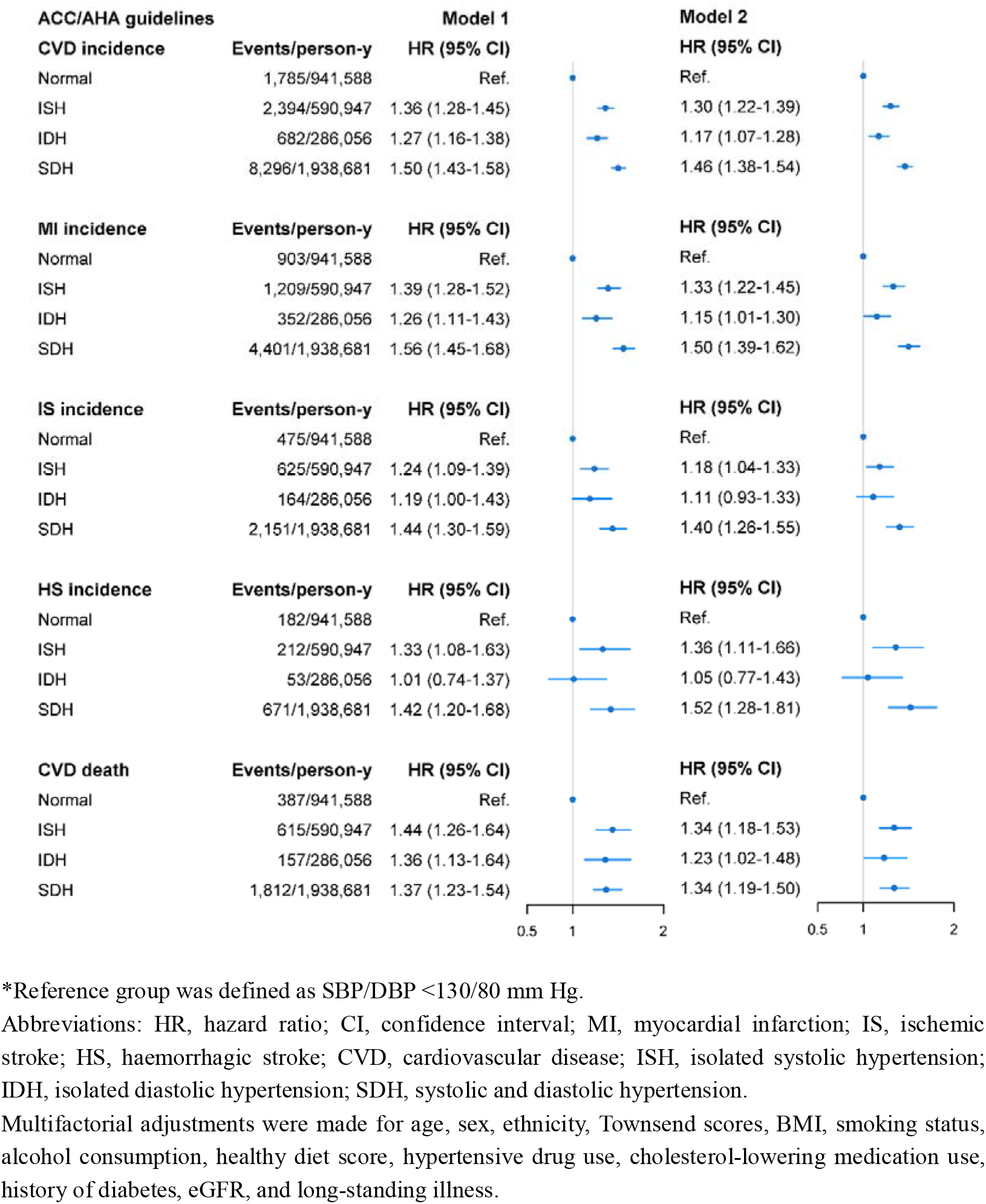
Multivariate-adjusted HRs (95% CIs) for the risk of CVD by hypertension subtype according to the ACC/AHA definition, after re-defining the reference group*.

